# Impacts of Morally Distressing Experiences on the Mental Health of Canadian Health Care Workers During the COVID-19 Pandemic

**DOI:** 10.1101/2021.04.06.21254800

**Authors:** Rachel A. Plouffe, Anthony Nazarov, Callista A. Forchuk, Dominic Gargala, Erisa Deda, Tri Le, Jesse Bourret-Gheysen, Brittni Jackson, Vanessa Soares, Fardous Hosseiny, Patrick Smith, Maya Roth, Arlene G. MacDougall, Michelle Marlborough, Rakesh Jetly, Alexandra Heber, Joy Albuquerque, Ruth Lanius, Ken Balderson, Gabrielle Dupuis, Viraj Mehta, J. Don Richardson

## Abstract

**Objective:** Research is urgently needed to understand health care workers’ (HCWs’) experiences of moral-ethical dilemmas encountered throughout the COVID-19 pandemic, and their associations with organizational perceptions and personal well-being. The purpose of this research is to evaluate associations between workplace experiences during COVID-19, moral distress, and the psychological well-being of Canadian HCWs.

**Method:** A total of 1383 French- and English-speaking Canadian HCWs employed during the COVID-19 pandemic were recruited to participate in an online survey. Participants completed measures reflecting moral distress, perceptions of organizational response to the pandemic, burnout, and symptoms of psychological disorders, including depression, anxiety, and posttraumatic stress disorder.

**Results:** Structural equation modeling showed that when organizational predictors were considered together, resource adequacy, positive work life impact, and ethical work environment negatively predicted severity of moral distress, whereas COVID-19 risk perception positively predicted severity of moral distress. Moral distress also significantly and positively predicted symptoms of depression, anxiety, PTSD, and burnout.

**Conclusions:** Our findings highlight an urgent need for HCW organizations to implement strategies designed to prevent long-term moral and psychological distress within the workplace. Ensuring availability of adequate resources, reducing HCW risk of contracting COVID-19, providing organizational support regarding individual priorities, and upholding ethical considerations are crucial to reducing severity of moral distress in HCWs.

---

In early 2020, the World Health Organization declared global pandemic due to the novel severe acute respiratory syndrome known as SARS-CoV-2 (COVID-19). As of March 2021, the COVID-19 pandemic has caused over 2.6 million deaths across 219 countries worldwide.^1^ In addition to high levels of morbidity and mortality,^2^ this pandemic has since resulted in devastating impacts on individuals’ psychological well-being.^3^

Since the onset of the pandemic, health care workers (HCWs) have faced unprecedented situations involving potentially life-altering moral-ethical decisions, including rationing of limited medical resources (e.g., beds, ventilators) to patients who are equally in need,^4^ and the possibility of ending life for some patients to allocate resources to those with fewer comorbidities.^5^ These complex moral-ethical decisions may have detrimental consequences for patients and HCWs alike, including reductions in quality of care^6^ and HCW moral distress.^7^ The overarching purpose of this study is to evaluate the associations between workplace experiences during COVID-19, moral distress, and the psychological well-being of HCWs.

## COVID-19 and Moral Distress in Health Care Workers

HCWs may become increasingly vulnerable to experiencing high severity of moral distress as a result of the moral-ethical challenges encountered throughout the COVID-19 pandemic.^7, 8^ Moral distress occurs when an individual is constrained to behave in a way that they perceive as morally or ethically inappropriate, and as such, they are unable to act in accordance with their core values.^9, 10^ For example, moral distress may occur if HCWs must attend to patients without typical recommended personal protective equipment (PPE), risking infection to self, to patients, or to family and community members, and are thus forced to compromise their typical standard of care.^8, 11-13^

Moral distress has been related to poor psychiatric outcomes in HCWs, including symptoms of depression and anxiety, even prior to the pandemic.^14-16^ Furthermore, throughout the pandemic, inconsistent and delayed guidance from government and workplace leaders may be associated with feelings of distrust and betrayal that could lead to doubts about the integrity of trusted figures and values.^12^ It is possible that this distrust could call into question the moral soundness of organizational policies and associated job requirements under the pandemic, further increasing the likelihood of moral distress.

## Moral Distress and Associations with Organizational Variables

HCWs’ experiences with moral-ethical dilemmas may be associated with adverse professional outcomes and perceptions of organizational environments, collectively referred to here as organizational variables or factors. Past empirical research has demonstrated that for nurses and physicians, high severity of moral distress were associated with negative professional outcomes, including increases in burnout, secondary traumatic stress, and intent to leave one’s professional position, as well as decreases in job satisfaction and perceptions of an ethical work environment.^14, 16-21^ In addition to these organizational factors, nurses and physicians have also reported concerns regarding quality and safety of care delivered to patients,^22-25^ which results in increased severity of moral distress.^25^ Although associations between workplace experiences and moral distress have not been empirically tested in HCWs during the COVID-19 pandemic, it is plausible to hypothesize that these findings would generalize across physicians, nurses, and other HCWs worldwide.

To best prevent long-term moral and psychological distress and to ensure that workers can optimally provide health services, research is urgently needed to understand experiences of workplace moral-ethical dilemmas throughout the COVID-19 pandemic, and their associations with HCWs’ organizational perceptions and personal well-being.

## Aims of the Study

The first objective of this study is to evaluate the effect of organizational variables on frequencies/severity of moral distress during the COVID-19 pandemic within a Canadian HCW sample. Based on past findings,^17, 19, 20^ we hypothesize that perceptions of COVID-19 risk in an organizational setting will positively predict severity of moral distress during the COVID-19 pandemic. Conversely, we predict that perceptions of an ethical work environment, adequacy of resources, leadership, and positive work life impact will negatively predict severity of moral distress.

Next, we aim to assess the effect of frequencies/severity of moral distress during the COVID-19 pandemic on symptoms of psychiatric disorders. Based on past findings,^14-16^ we predict that severity of moral distress will positively predict symptoms of depression, anxiety, posttraumatic stress disorder (PTSD), and burnout. This study is the first to empirically examine the relations between moral distress, organizational factors, and symptoms of psychiatric disorders among participants across health care professions in the context of the COVID-19 pandemic.

## Method

### Participants

Participants included 1383 English- or French-speaking HCWs employed across Canada. We defined HCWs as those “who provide health treatment and advice based on formal training and experience, or who work to directly support those providers in a clinical setting.” HCWs were eligible to participate if they were at least 18 years of age and currently or previously employed as a HCW in Canada between the start of the pandemic and the baseline end date.

### Measures

#### Moral Distress

Moral distress was evaluated using the Measure of Moral Distress for Healthcare Professionals (MMD-HP).^26^ The MMD-HP comprises 27 items measured on 4-point scales designed to assess the frequency (0=*never*, 4=*very frequently*) and distress severity (0=*none*, 4=*very distressing*) associated with dilemmas that may cause moral distress in the workplace. Composite scores were calculated by multiplying participants’ frequency scores by their distress scores.^26^ Higher scores represent higher severity of moral distress. Past research has supported the reliability and validity of the MMD-HP (e.g., α = .93).^26^

#### Burnout

The Expanded Well-Being Index (WBI)^27^ was used to assess severity of HCW burnout throughout the past month. The WBI consists of seven dichotomous (*yes/no*) items, and two additional items that evaluate how meaningful the individual’s work is to them (7-point scale from *very strongly disagree* to *very strongly agree*) and work-life balance (5-point scale from *strongly agree* to *strongly disagree*). Scores were calculated in accordance with scoring guidelines, ranging from −2 to 9, with higher scores indicating more burnout.^27^ Past empirical research has supported the validity of the WBI.^28, 29^

#### Organizational Response to the Pandemic

We assessed HCWs’ perceptions of their organizations’ responses to the COVID-19 pandemic using an adapted version of the Pandemic Experiences and Perceptions Survey (PEPS).^30^ The PEPS measures organizational response to the pandemic across five domains: Disruption (i.e., to workflow; 1=*no effect at all* to 5=*completely dominated the work*), Resource Adequacy (e.g., PPE; 1=*completely inadequate* to 5=*completely adequate*), COVID-19 Risk Perception (*1=no risk at all* to 7=*life threatening risk*), Positive Work Life Impact (e.g., work hours, social support; 1=*strongly disagree* to 5=*strongly agree*), and Leadership (i.e., supervisor/management; 1=*not at all* to 5=*frequently, if not always*). We included all domains except for Disruption. Mean scores on each subscale were calculated to create four separate scores, with higher scores indicating higher levels of resource adequacy, risk perception, more positive work life impact, and stronger leadership. The validity of the PEPS has been supported in recent research.^31^

#### Ethical Work Environment

We used the 20-item Ethics Environment Questionnaire (EEQ)^32^ to measure health care workers’ perceptions about ethics in their organizations on a 5-point scale. (1*=strongly disagree* to 5=*strongly agree*). We calculated participant mean scores on the EEQ, such that higher scores indicated a more ethical work environment. Past research has supported the reliability and validity of the EEQ (e.g., α = .93).^18^

#### Depression

HCWs’ depression symptom severity was assessed using the Patient Health Questionnaire-9 (PHQ-9).^33^ Participants responded to items on a scale ranging from 0=*not at all* to 3=*nearly every day* about their depressive symptoms throughout the past 14 days. Item responses were summed, such that higher scores reflect greater depression symptom severity. The PHQ-9 has demonstrated strong reliability and validity in past research (e.g., α = .89).^34^

#### Anxiety

To assess anxiety symptoms among HCWs, we used the 7-item GAD-7.^35^ Participants responded to items on a scale ranging from 0=*not at all* to 3=*nearly every day* about their anxiety symptoms throughout the past 14 days. Item responses were summed for a total anxiety score, with higher scores representing more severe anxiety. Past research has supported the reliability and validity of the GAD-7 (e.g., α = .89).^36^

#### Posttraumatic Stress Disorder

The 20-item PTSD Checklist for the DSM-5 (PCL-5)^37^ was used to measure PTSD symptom severity over the past month. Participants endorsed items on a 5-point scale ranging from 0=*not at all* to 4=*extremely*. Item responses were summed to create a total score, with higher scores reflecting greater PTSD symptom severity. Empirical research supports the reliability and validity of the PCL-5 (e.g., α = .94)□.^37^

### Procedure

Data presented here were drawn from a bilingual (i.e., English and French) survey exploring longitudinal changes in psychological functioning of HCWs during the COVID-19 pandemic. This research was approved by the ethical review board at Western University, Ontario, Canada. HCWs were recruited to participate via word of mouth, social media and online advertisements, participant recruitment websites, and media releases through Lawson Health Research Institute between June 26, 2020 and December 29, 2020. Interested participants were directed to the survey-hosting platform, Research Electronic Data Capture (REDCap), where they read a letter of information and provided informed consent to volunteer in the study.

Participants were provided with a choice to complete a long-form (approximately 25-minuteduration) or short-form (approximately 10-minute duration) of the study. They then completed a series of demographic items and surveys related to their organizational experiences, burnout, and psychiatric symptoms. Participants completed follow-up surveys every three months; results presented here are based on baseline data collection.

### Data Analytic Strategy

Participant total scores were created for those who had completed at least 80% of the measures’ items except for the PEPS subscales. For the PEPS, participant scores were calculated for those who completed at least 50% due to the small number of items. Preliminary analyses, including descriptive statistics and bivariate correlations were calculated for all study variables using SPSS Version 26.0.^38^ Next, we tested predictive models to assess the associations between moral distress and organizational variables and psychiatric sequelae using structural equation modeling (SEM) in MPlus Version 8^39^. We used the maximum likelihood robust estimator to correct standard errors for data non-normality, and the default full-information maximum likelihood to estimate missing data. To ensure that our sample size was adequate with consideration for model parameters,^40^ we conducted two separate SEMs.

In the first model, we regressed the indicator variable, moral distress, onto latent variables associated with each subscale of the PEPS, as well as latent EEQ. We used parcels as indicators for the EEQ, as this measure comprises 20 unidimensional items, and parceling is recommended to stabilize parameter estimates for measures with a large number of items.^41^ We used an indicator variable to represent moral distress because the MMD-HP aligns more with a formative (cf. reflective) model of measurement, and as such, it is not appropriate to specify that the exposure indicators occur as a result of latent moral distress.^42^

In the second model, we regressed outcome variables, including symptoms of anxiety, depression, PTSD, and burnout on moral distress. We used PCL-5 symptom cluster subscores as indicator variables for PTSD symptoms. Burnout was classified as an indicator variable, as its final two items were measured on Likert scales in contrast to the remaining dichotomous items. Model fit was established using cut-off values recommended by Hu and Bentler.^43^

## Results

### Sample Characteristics

The top five occupations in the current study included nursing (42.8%), community health/personal support worker (10.6%), physician (4.8%), paramedic (4.2%), and social worker (3.0%). Of those enrolled in the study, *n* = 377 indicated they were working part-time or casual, and *n* = 1006 were working full-time. A total of 804 HCWs were directly involved in clinical activities including diagnosis, treatment, or provision of direct care to patients with elevated temperature or confirmed COVID-19 in the month preceding participation. Additional demographic information and more detailed occupational information is presented in Table 1.

**Table 1.** Demographic and Organization Information

| Variable | Frequency | Percentage (%) |
| --- | --- | --- |
| Age (years) |  |  |
| ≤25 | 40 | 2.9 |
| 26-40 | 447 | 32.3 |
| 41-60 | 548 | 39.6 |
| >60 | 65 | 4.7 |
| Prefer not to answer | 4 | 0.3 |
| Missing | 279 | 20.2 |
| Gender |  |  |
| Women | 986 | 71.3 |
| Men | 96 | 6.9 |
| Other | 7 | 0.5 |
| Prefer not to answer | 14 | 1.0 |
| Missing | 280 | 20.2 |
| Marital status |  |  |
| Single | 228 | 16.5 |
| Married/common law | 718 | 51.9 |
| Separated | 46 | 3.3 |
| Divorced | 77 | 5.6 |
| Widowed | 10 | 0.7 |
| Prefer not to answer | 27 | 2.0 |
| Missing | 277 | 20.0 |
| Province/territory |  |  |
| Prince Edward Island | 4 | 0.3 |
| Northwest Territories | 5 | 0.4 |
| Newfoundland | 7 | 0.5 |
| New Brunswick | 14 | 1.0 |
| Nova Scotia | 21 | 1.5 |
| Quebec | 34 | 2.5 |
| Saskatchewan | 35 | 2.5 |
| Manitoba | 48 | 3.5 |
| British Columbia | 95 | 6.9 |
| Alberta | 129 | 9.3 |
| Ontario | 441 | 31.9 |
| Missing | 550 | 39.8 |
| Highest level of education |  |  |
| Secondary or lower | 29 | 2.1 |
| Post-secondary or higher | 1077 | 77.9 |
| Missing | 277 | 20.0 |
| Primary job function |  |  |
| Administration | 89 | 6.4 |
| Outreach | 13 | 0.9 |
| Research | 15 | 1.1 |
| Direct client/patient care | 1186 | 85.8 |
| Other | 65 | 4.7 |
| Missing | 15 | 1.1 |
| Number of years as a health care worker |  |  |
| <6 months | 9 | 0.7 |
| 6 months to 1 year | 20 | 1.4 |
| 1 to 5 years | 256 | 18.5 |
| 6 to 10 years | 284 | 20.5 |
| 11+ years | 809 | 58.5 |
| Missing | 5 | 0.4 |
| Percentage direct patient care |  |  |
| No direct patient care | 77 | 5.4 |
| 1-24% | 72 | 5.2 |
| 25-50% | 79 | 5.7 |
| 51-74% | 152 | 11.0 |
| 75-100% | 997 | 72.1 |
| Missing | 6 | 0.4 |
| Workplace setting |  |  |
| Private practice | 118 | 8.5 |
| Hospital/community health centre | 940 | 68.0 |
| Other | 323 | 23.4 |
| Missing | 2 | 0.1 |

### Preliminary Analyses

Descriptive statistics, Cronbach’s alpha values, and bivariate correlations are reported in Table 2. All bivariate correlations between study variables were statistically significant (*p*<.001) with effect sizes ranging in magnitude from small to large.

**Table 2.**
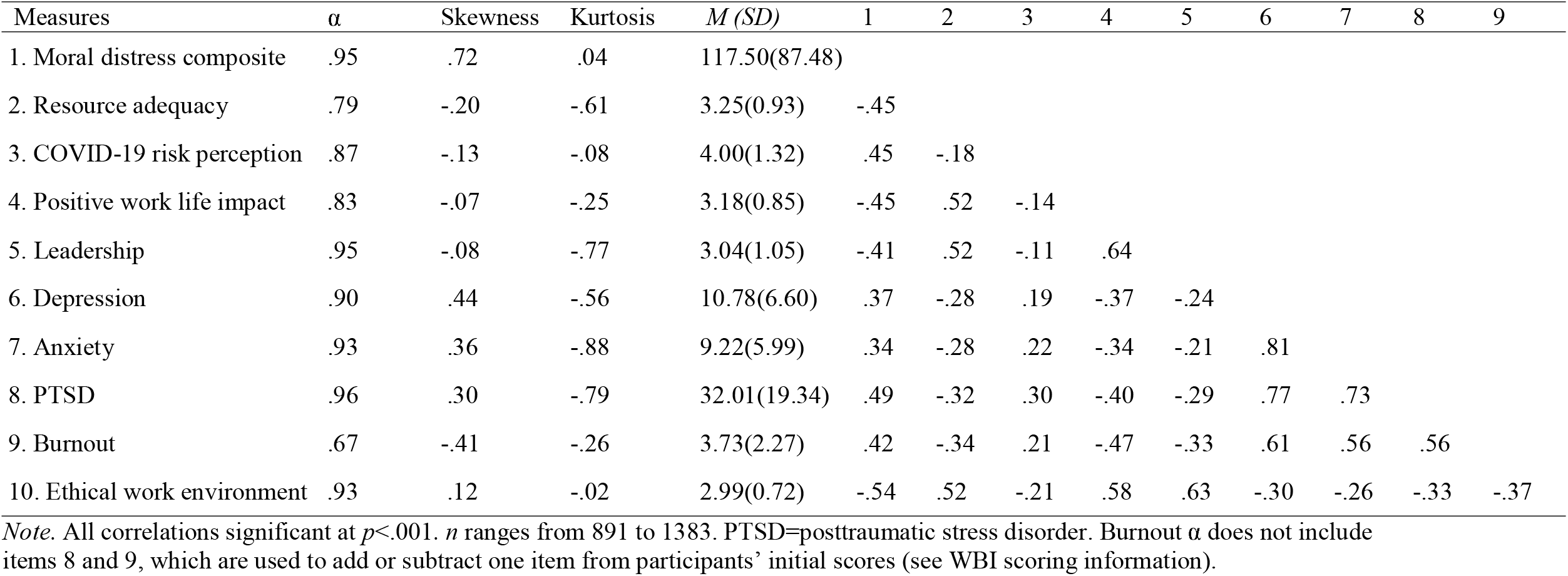
Descriptive Statistics and Bivariate Correlations

### Structural Equation Models

The first SEM regressing moral distress on PEPS variables and ethical work environment demonstrated poor fit (see Model 1 in Table 3). Upon closer inspection, modification indices demonstrated that correlating errors on the PEPS Leadership subscale would improve model fit, as item pairs reflected identical content and were endorsed separately for organizational management and immediate supervisor. To account for these identical item pairs, we conducted two additional structural models: Model 2) two separate latent variables representing Organizational Management Leadership and Immediate Supervisor Leadership, and Model 3) correlated errors between seven Leadership item pairs based on modification indices. Although model fit improved slightly (Table 3), modification indices showed that errors should again be correlated across the two latent Leadership variables due to content overlap. In addition, correlating errors without a priori theoretical rationale can inflate model fit and reduce potential for replication.^44, 45^ We believe that the psychometric properties of the Leadership subscale should be revisited prior to its use in research, and therefore, we did not include Leadership in the final model.

**Table 3.** Structural Equation Model Fit Indices

| Model | $\chi^2$ (df) | CFI | TLI | RMSEA | RMSEA 90% CI |
| --- | --- | --- | --- | --- | --- |
| Moral distress as outcome |  |  |  |  |  |
| Model 1: All outcomes | 3998.871*(420) | .840 | .823 | .078* | .076, .081 |
| Model 2: Two leadership latent variables | 2824.257*(414) | .892 | .879 | .065* | .063, .067 |
| Model 3: Leadership correlated errors | 2524.400*(413) | .906 | .894 | .061* | .059, .063 |
| Model 4: Final model without leadership | 998.231*(180) | .925 | .913 | .057* | .054, .061 |
| Moral distress as predictor | 1247.556*(201) | .921 | .909 | .067* | .063, .070 |

*Note.* CFI = comparative fit index. TLI = Tucker-Lewis index. RMSEA = root mean square error of approximation. CI = confidence interval. \* $p < .001$ .

The SEM without including the Leadership subscale included showed strong fit (see Model 4 in Table 3). As shown in Figure 1, resource adequacy, positive work life impact, and ethical work environment negatively predicted severity of moral distress, whereas COVID-19 risk perception positively predicted severity of moral distress. Finally, the SEM regressing symptoms of depression, anxiety, PTSD, and burnout on moral distress fit the data well (see Table 3). As shown in Figure 2, moral distress significantly and positively predicted symptoms of depression, anxiety, PTSD, and burnout.

**Figure 1.**
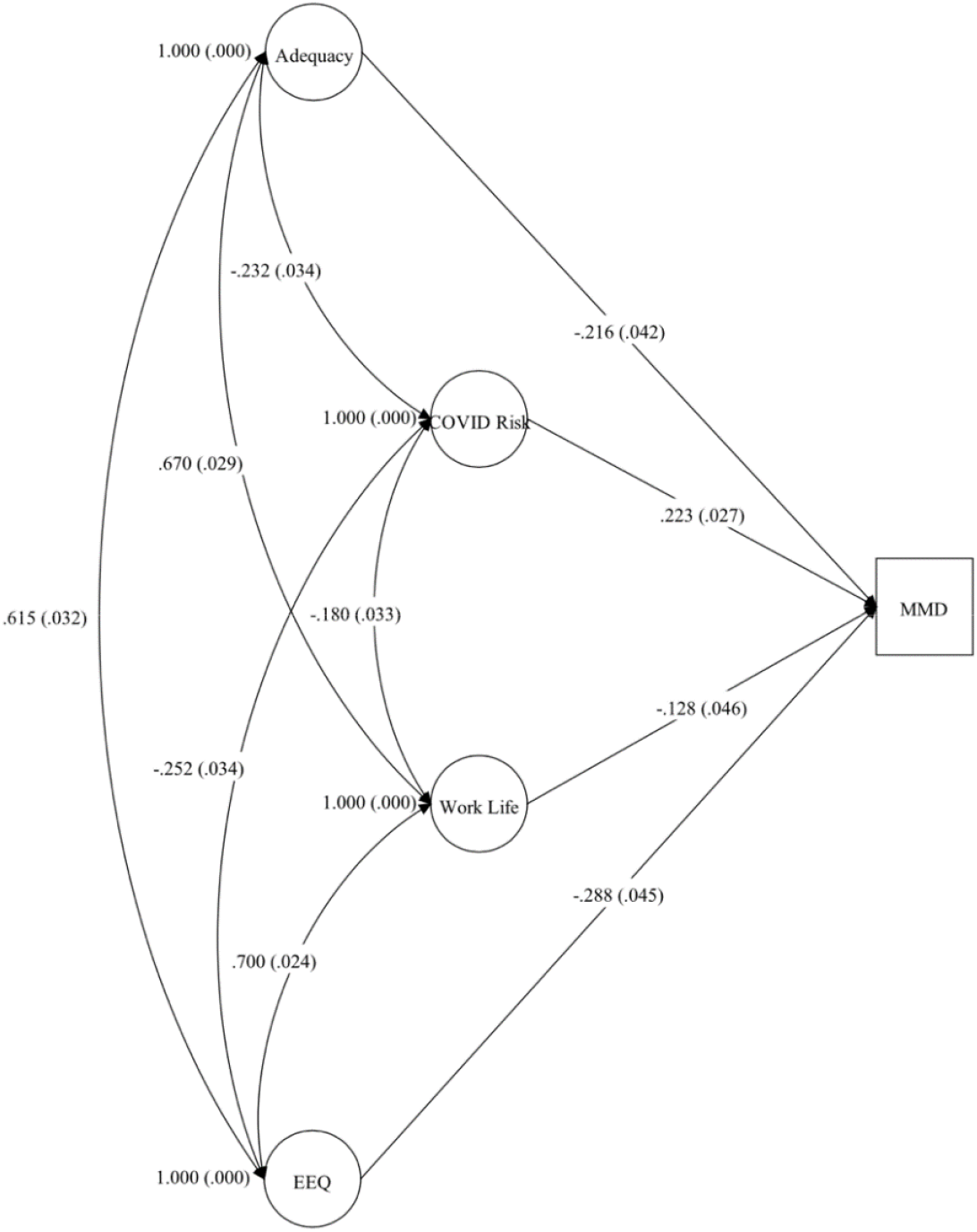
Moral Distress Regressed on Organizational Variables and Ethical Work Environment Without Leadership: Standardized Path Coefficients and Latent Variable Correlations *Note. n* = 1383. MMD = Moral distress composite. EEQ = Ethics Environment Questionnaire. Adequacy = Resource Adequacy. Coefficients are standardized. Standard error in brackets. All path coefficients and latent variable correlations significant at *p* < .01.

**Figure 2.**
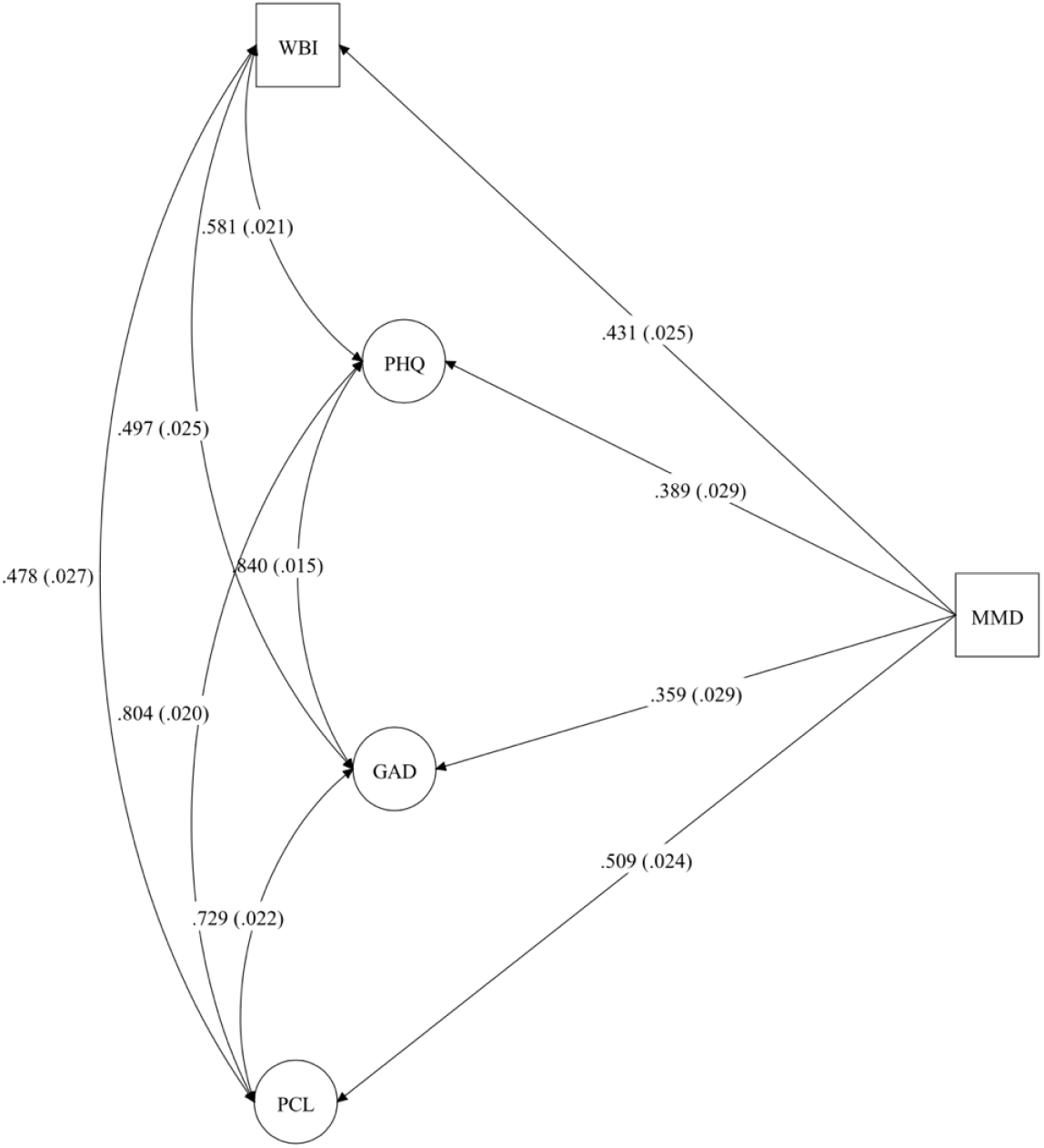
Burnout, Depression, Anxiety, and PTSD Symptoms Regressed on Moral Distress: Standardized Path Coefficients and Latent Variable Correlations *Note. n* = 1175. MMD = Moral distress composite. WBI = Well-Being Index. PHQ = Patient Health Questionnaire (depression symptoms). GAD = GAD-7 (anxiety symptoms). PCL = PTSD Checklist for the DSM-5. Coefficients are standardized. Standard error in brackets. All path coefficients and latent variable correlations significant at *p* < .001.

## Discussion

As expected, bivariate correlations showed that COVID-19-related organizational variables, including workplace resource adequacy, positive work life impact, managerial and organizational leadership, and perceptions of an ethical work environment are negatively associated with moral distress composite scores. On the other hand, moral distress was positively related to scores on COVID-19 risk perception. These findings are consistent with literature indicating that an organization’s response to the COVID-19 pandemic plays a strong role in HCW exposure and distress associated with moral-ethical dilemmas.^46, 47^ Specifically, issues encountered throughout (and even prior to) the pandemic, such as rationing of limited PPE and additional resources required to treat patients,^7, 48, 49^ lack of organizational leadership,^16, 46^ and potential contact with the virus^47^ are all factors that contribute to the development and maintenance of moral distress in HCWs. Furthermore, when HCW organizations integrate core ethical values in their processes and strategies, this has been associated with reduced severity of moral distress, even prior to the COVID-19 pandemic.^18^

These results were corroborated using SEM. Specifically, when all organizational variables were considered together, workplace resource adequacy, positive work life impact, and perceptions of an ethical work environment negatively predicted severity on moral distress, whereas COVID-19 risk perceptions positively predicted severity on moral distress.

Interestingly, managerial and supervisor leadership was not a significant predictor of moral distress when all variables were included in the model. Upon inspection of the modification indices, it was evident that there was a surplus of shared error variance between item pairs reflecting managerial and supervisor leadership due to their identical wording. We opted to interpret a model without Leadership included, as this measure should be refined prior to future use. Similar to the model with Leadership included, each of the organizational variables significantly predicted severity of moral distress. Overall, these findings suggest that organizational responses to COVID-19 play a role in HCWs’ experiences of moral-ethical dilemmas and distress associated with such dilemmas.

As hypothesized, bivariate relations between moral distress and psychiatric outcomes, including burnout, depression, anxiety, and PTSD symptoms were significant and medium in magnitude based on Cohen’s guidelines.^50^ When the variables were entered into an SEM, similar results emerged, such that moral distress predicted levels of latent burnout, depression, anxiety, and PTSD symptoms with medium-to-large effect sizes. These findings support the notion that when HCWs engage in activities that they perceive to be morally or ethically conflicting, such as rationing scant PPE and risking infection to self and others, individuals may perceive themselves as failing to uphold their core personal and professional values and roles as caregivers to patients.^8, 11, 17^ This, in turn, leads to feelings and symptoms of guilt, shame, anger, anxiety, traumatic stress, burnout, and depression.^8, 14-17^

Overall, our findings highlight an urgent need for HCW organizations to implement strategies designed to prevent moral and psychological distress within the workplace. Specifically, our results indicated that ensuring the perceived availability of adequate resources (e.g., PPE, ventilators, medications, staffing numbers), reducing HCW risk of contracting COVID-19, providing organizational support regarding decisions and individual priorities, and upholding ethical considerations are crucial to reducing severity of moral distress in HCWs.

## Limitations and Future Directions

This study is not without limitations. Although we sampled a large number of HCWs from each Canadian province/territories, our sample is not representative of the greater Canadian HCW population (e.g., geographical, occupational, gender). In addition, our sample comprised mostly women. However, this is not surprising, as the most common HCW occupation in our sample comprises mainly women (nursing). Future research should ensure accurate representation of the Canadian HCW population using stratified sampling strategies, accounting for non-response bias, and exploring gender-specific effects.

The data presented here were cross-sectional in nature, which limits the conclusions that can be drawn from our findings. Longitudinal data reflecting moral distress and well-being to test mediation hypotheses, including whether moral distress mediates the associations between organizational variables and psychiatric outcomes measured, may elucidate the causal relationships among these domains. Finally, more research exploring the nuances of moral distress in HCWs (e.g., potential gender and occupational group differences, longitudinal trajectories) will inform the optimal development of resilience interventions aimed at the individual, group, institutional, and government levels.

## Concluding Remarks

Overall, our findings provide insight into the moral-ethical dilemmas and the major detrimental effects these dilemmas have on HCWs’ psychological well-being. To best prevent long-term moral and psychological distress and to provide high quality care to patients, HCWs and their organizations, communities, and governments must work together and communicate standardized decision-making processes (e.g., regarding PPE distribution) effectively to ensure that core ethical principles are upheld and that the mental and physical well-being of HCWs are maintained.

## Data Availability

Study data are available from the corresponding author, RP, upon reasonable request.

## Acknowledgments/Funding

This project was funded through the support of the MacDonald Franklin OSI Research Centre by the St. Joseph’s Health Care Foundation (London, Ontario, Canada) and through a partnership with the Centre of Excellence for PTSD (Ottawa, Ontario, Canada).

## References

1. World Health Organization. WHO Coronavirus (COVID-19) Dashboard. Updated March 08, 2021. Accessed March 08, 2021, 2021. https://covid19.who.int/?gclid=Cj0KCQiAs5eCBhCBARIsAEhk4r4dsuP522kuirJJazxnGZ9DEMcaEN4PjJr796Yo0GkENqQ_DcUuAn0aAjwrEALw_wcB

2. Grech V. Unknown unknowns–COVID-19 and potential global mortality. Early human development. 2020;144:105026.

3. Serafini G, Parmigiani B, Amerio A, Aguglia A, Sher L, Amore M. The psychological impact of COVID-19 on the mental health in the general population. QJM: An International Journal of Medicine. 2020;113(8):531–537.

4. Greenberg N. Mental health of health-care workers in the COVID-19 era. Nature Reviews Nephrology. 2020:1-2.

5. Williamson V, Murphy D, Greenberg N. COVID-19 and experiences of moral injury in front-line key workers. Oxford University Press UK; 2020.

6. Kok N, Hoedemaekers A, van der Hoeven H, Zegers M, van Gurp J. Recognizing and supporting morally injured ICU professionals during the COVID-19 pandemic. Intensive Care Medicine. 2020;46:1653–1654.

7. Khoo EJ, Lantos JD. Lessons learned from the COVIDL19 pandemic. Acta Paediatrica. 2020;109(7):1323–1325.

8. PTSD PACfPMHatCCoE. PTSD PACfPMHatCCoE, ed. Moral Stress Amongst Healthcare Workers During COVID-19: A Guide to Moral Injury. 2020. https://www.moralinjuryguide.ca/wp-content/uploads/2020/07/Moral-Injury-Guide.pdf

9. Hamric AB. A case study of moral distress. Journal of Hospice & Palliative Nursing. 2014;16(8):457–463.

10. Jameton A. Dilemmas of moral distress: moral responsibility and nursing practice. AWHONN’s clinical issues in perinatal and women’s health nursing. 1993;4(4):542–551.

11. Binkley CE, Kemp DS. Ethical rationing of personal protective equipment to minimize moral residue during the COVID-19 pandemic. Journal of the American College of Surgeons. 2020;230(6):1111–1113.

12. Hossain F. Moral distress among healthcare providers and mistrust among patients during COVIDL19 in Bangladesh. Developing world bioethics. 2020;

13. Ranney ML, Griffeth V, Jha AK. Critical supply shortages—the need for ventilators and personal protective equipment during the Covid-19 pandemic. New England Journal of Medicine. 2020;382(18):e41.

14. Christodoulou-Fella M, Middleton N, Papathanassoglou ED, Karanikola MN. Exploration of the association between nurses’ moral distress and secondary traumatic stress syndrome: implications for patient safety in mental health services. BioMed research international. 2017;2017

15. Colville G, Dawson D, Rabinthiran S, Chaudry-Daley Z, Perkins-Porras L. A survey of moral distress in staff working in intensive care in the UK. Journal of the Intensive Care Society. 2019;20(3):196–203.

16. Lamiani G, Borghi L, Argentero P. When healthcare professionals cannot do the right thing: A systematic review of moral distress and its correlates. Journal of health psychology. 2017;22(1):51–67.

17. Austin CL, Saylor R, Finley PJ. Moral distress in physicians and nurses: Impact on professional quality of life and turnover. Psychological Trauma: Theory, Research, Practice, and Policy. 2017;9(4):399.

18. Corley MC, Minick P, Elswick R, Jacobs M. Nurse moral distress and ethical work environment. Nursing ethics. 2005;12(4):381–390.

19. de Veer AJ, Francke AL, Struijs A, Willems DL. Determinants of moral distress in daily nursing practice: A cross sectional correlational questionnaire survey. International journal of nursing studies. 2013;50(1):100–108.

20. Fumis RRL, Amarante GAJ, de Fátima Nascimento A, Junior JMV. Moral distress and its contribution to the development of burnout syndrome among critical care providers. Annals of intensive care. 2017;7(1):1–8.

21. Pauly B, Varcoe C, Storch J, Newton L. Registered nurses’ perceptions of moral distress and ethical climate. Nursing ethics. 2009;16(5):561–573.

22. Lerkiatbundit S, Borry P. Moral distress part I: critical literature review on definition, magnitude, antecedents and consequences. Thai J Pharm Pract. 2009;1(1):1–10.

23. Mrayyan MT, Hamaideh SH. Clinical errors, nursing shortage and moral distress: The situation in Jordan. Journal of Research in Nursing. 2009;14(4):319–330.

24. Wilkinson JM. Moral distress in nursing practice: experience and effect. Wiley Online Library; 1987:16–29.

25. Wolf LA, Perhats C, Delao AM, Moon MD, Clark PR, Zavotsky KE. “It’s a burden you carry”: describing moral distress in emergency nursing. Journal of Emergency Nursing. 2016;42(1):37–46.

26. Epstein EG, Whitehead PB, Prompahakul C, Thacker LR, Hamric AB. Enhancing understanding of moral distress: the measure of moral distress for health care professionals. AJOB empirical bioethics. 2019;10(2):113–124.

27. Dyrbye LN, Satele D, Shanafelt T. Ability of a 9-item well-being index to identify distress and stratify quality of life in US workers. Journal of occupational and environmental medicine. 2016;58(8):810–817.

28. Dyrbye LN, Satele D, Sloan J, Shanafelt TD. Ability of the physician well-being index to identify residents in distress. Journal of graduate medical education. 2014;6(1):78.

29. Dyrbye LN, Schwartz A, Downing SM, Szydlo DW, Sloan JA, Shanafelt TD. Efficacy of a brief screening tool to identify medical students in distress. Academic Medicine. 2011;86(7):907–914.

30. Leiter M. The Pandemic Experiences and Perceptions Survey (PEPS). Group report. 2020:1–57.

31. AlMulla M. A measurement of radiographers pandemic experiences & perceptions survey (PEPS) during the coronavirus pandemic in Kuwait. 2020;

32. McDaniel C. Development and psychometric properties of the ethics environment questionnaire. Medical Care. 1997;35(9):901–914.

33. Spitzer RL, Kroenke K, Linzer M, et al. Health-related quality of life in primary care patients with mental disorders: results from the PRIME-MD 1000 study. Jama. 1995;274(19):1511–1517.

34. Kroenke K, Spitzer RL, Williams JB. The PHQL9: validity of a brief depression severity measure. Journal of general internal medicine. 2001;16(9):606–613.

35. Spitzer RL, Kroenke K, Williams JB, Löwe B. A brief measure for assessing generalized anxiety disorder: the GAD-7. Archives of internal medicine. 2006;166(10):1092–1097.

36. Löwe B, Decker O, Müller S, et al. Validation and standardization of the Generalized Anxiety Disorder Screener (GAD-7) in the general population. Medical care. 2008:266-274.

37. Blevins CA, Weathers FW, Davis MT, Witte TK, Domino JL. The posttraumatic stress disorder checklist for DSML5 (PCLL5): Development and initial psychometric evaluation. Journal of traumatic stress. 2015;28(6):489–498.

38. IBM SPSS Statistics for Windows. Version 26.0. IBM Corp.; 2019.

39. Muthén LK, Muthén BO. Mplus User’s Guide.. Eighth Edition. ed. Muthén & Muthén; 1998-2017.

40. Kline RB. Principles and practice of structural equation modeling. Guilford publications; 2015.

41. Matsunaga M. Item parceling in structural equation modeling: A primer. Communication Methods and Measures. 2008;2(4):260–293.

42. Bollen KA, Bauldry S. Three Cs in measurement models: causal indicators, composite indicators, and covariates. Psychological methods. 2011;16(3):265.

43. Hu Lt, Bentler PM. Cutoff criteria for fit indexes in covariance structure analysis: Conventional criteria versus new alternatives. Structural equation modeling: a multidisciplinary journal. 1999;6(1):1–55.

44. Hermida R. The problem of allowing correlated errors in structural equation modeling: concerns and considerations. Computational Methods in Social Sciences. 2015;3(1):5–17.

45. Landis RS, Edwards BD, Cortina JM. On the practice of allowing correlated residuals among indicators in structural equation models. 2008.

46. Dryden-Palmer K, Moore G, McNeil C, et al. Moral Distress of Clinicians in Canadian Pediatric and Neonatal ICUs. Pediatric critical care medicine. 2020;21(4):314-323. doi:10.1097/PCC.0000000000002189

47. Hlubocky FJ, Symington BE, McFarland DC, et al. Impact of the COVID-19 Pandemic on Oncologist Burnout, Emotional Well-Being, and Moral Distress: Considerations for the Cancer Organization’s Response for Readiness, Mitigation, and Resilience. JCO oncology practice. 2021:OP2000937-OP2000937. doi:10.1200/OP.20.00937

48. Borges LM, Barnes SM, Farnsworth JK, Bahraini NH, Brenner LA. A Commentary on Moral Injury Among Health Care Providers During the COVID-19 Pandemic. Psychological trauma. 2020;12(S1):S138-S140. doi:10.1037/tra0000698

49. Shanafelt T, Ripp J, Trockel M. Understanding and Addressing Sources of Anxiety Among Health Care Professionals During the COVID-19 Pandemic. JAMA. 2020;323(21):2133-2134. doi:10.1001/jama.2020.5893

50. Cohen J. A power primer. Psychological bulletin. 1992;112(1):155.

